# Yellow fever disease severity and endothelial dysfunction are associated with elevated serum levels of viral NS1 protein and syndecan-1

**DOI:** 10.1101/2023.06.29.23292053

**Authors:** Francielle T. G. de Sousa, Colin M. Warnes, Erika R. Manuli, Arash Ng, Luiz G. F. A. B. D’Elia Zanella, Yeh-Li Ho, Samhita Bhat, Camila M. Romano, P. Robert Beatty, Scott B. Biering, Esper G. Kallas, Ester C. Sabino, Eva Harris

## Abstract

Yellow fever virus (YFV) infections can cause severe disease manifestations, including hepatic injury, endothelial damage, coagulopathy, hemorrhage, systemic organ failure, and shock, and are associated with high mortality in humans. While nonstructural protein 1 (NS1) of the related dengue virus is implicated in contributing to vascular leak, little is known about the role of YFV NS1 in severe YF and mechanisms of vascular dysfunction in YFV infections. Here, using serum samples from qRT-PCR-confirmed YF patients with severe (n=39) or non-severe (n=18) disease in a well-defined hospital cohort in Brazil, plus samples from healthy uninfected controls (n=11), we investigated factors associated with disease severity. We developed a quantitative YFV NS1 capture ELISA and found significantly increased levels of NS1, as well as syndecan-1, a marker of vascular leak, in serum from severe YF as compared to non-severe YF or control groups. We also showed that hyperpermeability of endothelial cell monolayers treated with serum from severe YF patients was significantly higher compared to non-severe YF and control groups as measured by transendothelial electrical resistance (TEER). Further, we demonstrated that YFV NS1 induces shedding of syndecan-1 from the surface of human endothelial cells. Notably, YFV NS1 serum levels significantly correlated with syndecan-1 serum levels and TEER values. Syndecan-1 levels also significantly correlated with clinical laboratory parameters of disease severity, viral load, hospitalization, and death. In summary, this study points to a role for secreted NS1 in YF disease severity and provides evidence for endothelial dysfunction as a mechanism of YF pathogenesis in humans.

**Significance:** Yellow fever virus (YFV) infections cause a major global disease burden, and as such it is critical to identify clinical correlates of disease severity. Using clinical samples from our hospital cohort in Brazil, we show that YF disease severity is associated with increased serum levels of the viral nonstructural protein 1 (NS1) and soluble syndecan-1, a marker of vascular leak. This study extends the role of YFV NS1 in triggering endothelial dysfunction to human YF patients, previously demonstrated *in vitro* and in mouse models. Further, we developed a YFV NS1-capture ELISA that serves as a proof-of-concept for low-cost NS1-based diagnosis/prognosis tools for YF. Together, our data shows that YFV NS1 and endothelial dysfunction are important components of YF pathogenesis.

## Introduction

Yellow fever virus (YFV) is an arbovirus endemic to tropical areas of Central and South America and sub-Saharan Africa and the causative agent of yellow fever (YF) disease. YF is considered a reemerging disease, with increasing infections over the past 20 years, exemplified by epidemics in 2015 and 2016 in Angola and the Democratic Republic of Congo and in 2016-2019 in Brazil (1, 2). The clinical spectrum of YF in humans ranges from asymptomatic infection to mild illness to severe disease. Manifestations of severe disease include vasculopathy and organ impairment of the liver, kidneys, lungs, intestine, and brain, resulting in high reported case fatality rates (20-60%) (2).

YFV is a member of the *Flavivirus* genus of the *Flaviviridae* family, with a positive-sense RNA genome of approximately 11 kb that encodes three structural and seven nonstructural proteins (3). The *Flavivirus* nonstructural protein 1 (NS1) is secreted by infected cells and has been used as a serological diagnostic marker for dengue (4). Further, dengue virus (DENV) NS1 levels have been correlated with disease severity (5–7). We have recently shown that *Flavivirus* NS1 can directly cause vascular leak *in vitro* and in animal models in a tissue-specific manner that reflects the viral disease tropism (8, 9). Mechanisms of NS1-induced endothelial dysfunction include disruption of key barriers of endothelial integrity such as the glycocalyx (8) and intercellular junctional complexes (9, 10). Further, studies have demonstrated that soluble NS1, independently from the virus, can act on endothelial cells to facilitate viral dissemination and pathology (11, 12). In addition, DENV NS1 has been shown to increase circulating levels of glycocalyx components including sialic and hyaluronic acid, heparan sulfate, and syndecan-1 (SDC-1) *in vitro*, in animal models, and in clinical samples (8, 13–18). However, while YFV NS1 can trigger endothelial dysfunction *in vitro* and in animal models (8), a direct correlation of NS1 levels with severe YF disease in humans has not been demonstrated.

SDC-1, also known as CD138, is an extracellular matrix receptor and a member of the transmembrane heparan sulfate proteoglycan family. SDC-1 is involved in many cellular functions, including cell-cell and cell-matrix adhesion, and is highly expressed by epithelial, endothelial, and hematopoietic cells (19, 20). As a part of normal cell surface proteoglycan turnover, the ectodomain of SDC-1 is constitutively shed from the cell surface via proteolytic cleavage by metalloproteinases (21). Increased levels of soluble syndecan-1 (sSDC-1) have been detected in response to injury or infection, and sSDC-1 is often utilized as a marker for glycocalyx disruption. Therefore, sSDC-1 has been identified as a potential prognostic factor in cancer and systemic inflammatory and autoimmune diseases (19, 20, 22). In addition, sSDC-1 has been associated with severe manifestations of SARS-CoV-2 infection (23, 24), and elevated levels of sSDC-1 were found in patients with severe dengue due to the destruction of the glycocalyx associated with vascular leak (16, 25). Nonetheless, to date, a link between YF disease severity and glycocalyx disruption or increased serum levels of sSDC-1 has not yet been demonstrated.

In this study, we use a well-defined cohort of patients with YFV infection to examine the relationship between levels of YFV NS1, sSDC-1, and severe disease manifestations. We found significantly higher levels of YFV NS1 and sSDC-1 in sera of severe YF patients as compared to non-severe YF and control groups. YFV NS1 and sSDC-1 serum levels correlated with each other and with clinical laboratory signs of severe YF disease. We also found that treatment of endothelial monolayers with serum from individuals with severe YF induced significantly higher levels of endothelial permeability as compared to the non-severe YF and control groups. Further, YFV NS1 treatment of endothelial cells induced syndecan shedding, as observed by immunofluorescence assay. Together, these results provide evidence for a role of NS1 in YF disease and for endothelial glycocalyx disruption as a pathogenic mechanism in YFV infections in humans.

## Results

### Characteristics of study participants

In this study, we analyzed serum samples from individuals with suspected YF collected at the time of hospital admission during an observational cohort study initiated during the 2018 YF epidemic in São Paulo, Brazil, as previously described (2), and continued through the subsequent epidemic in 2019. Demographic characteristics, clinical manifestations, and laboratory data of study participants with RT-PCR-confirmed YF cases as well as healthy controls are provided in Table 1. We used the laboratory criteria (viral load, neutrophil count, aspartate transaminase [AST], creatinine, and indirect bilirubin [IB]) previously determined to predict mortality in YF patients from the same cohort (2), or death, to define severe cases. Table 1 shows that in addition to these criteria, severe YF cases also displayed significantly higher levels of alanine aminotransferase (ALT), total bilirubin (TB), and direct bilirubin (DB), as well as increased prothrombin time (PT/INR) and activated partial thromboplastin time (aPTT), when compared to the non-severe group. The majority of YF-positive study participants were male agriculture or forestry workers.

**Table 1:**
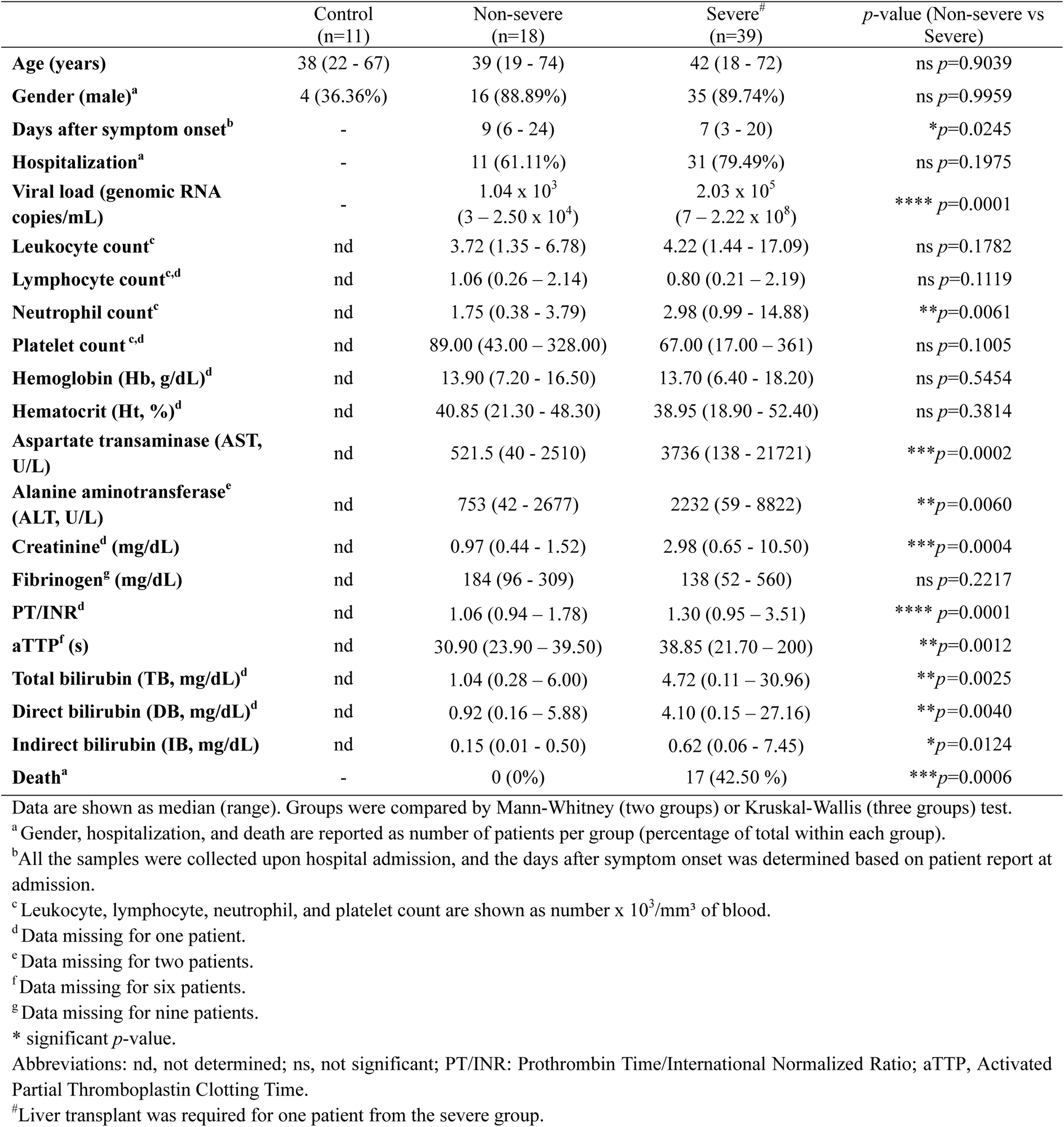
Demographic, clinical, and laboratory data of study participants.

### Development of a quantitative YFV NS1 capture ELISA for clinical samples

While YFV NS1 has been previously implicated as a direct trigger of endothelial dysfunction and vascular leak *in vitro* and in mouse models (8), the contribution of YFV NS1 to endothelial dysfunction in humans is unknown. To measure YFV NS1 levels in the serum of infected patients, we established a quantitative YFV NS1 capture ELISA using an in-house-produced mouse anti-YFV-NS1 monoclonal antibody (mAb). After testing different mAbs, YFJ19 (IgG1) used as the capture mAb and biotinylated YFJ19 as the detection mAb was determined to be the optimal combination, displaying the highest signal and specificity for YFV NS1 (Supplementary Fig. 1). Standard curves were generated via ELISA with recombinant YFV NS1 concentrations (0.24–2000 ng/mL) versus absorbance values (Fig. 1A), with no significant changes observed when human serum (1:10) was added (Fig. 1B), indicating that the assay could be performed with clinical samples. The analytic detection range was 2 to 500 ng/mL. YFJ19, as well as 2B7, a pan-flavivirus anti-NS1 mAb, was tested across a panel of 12 flavivirus NS1 proteins using both direct ELISA and Western blot methods. Results show that YFJ19, in contrast to 2B7, was specific to YFV NS1 (Fig. 1C, D, E). This specificity is important for assays detecting flavivirus NS1, especially because different flaviviruses can co-circulate in endemic areas such as Brazil.

**Fig. 1.**
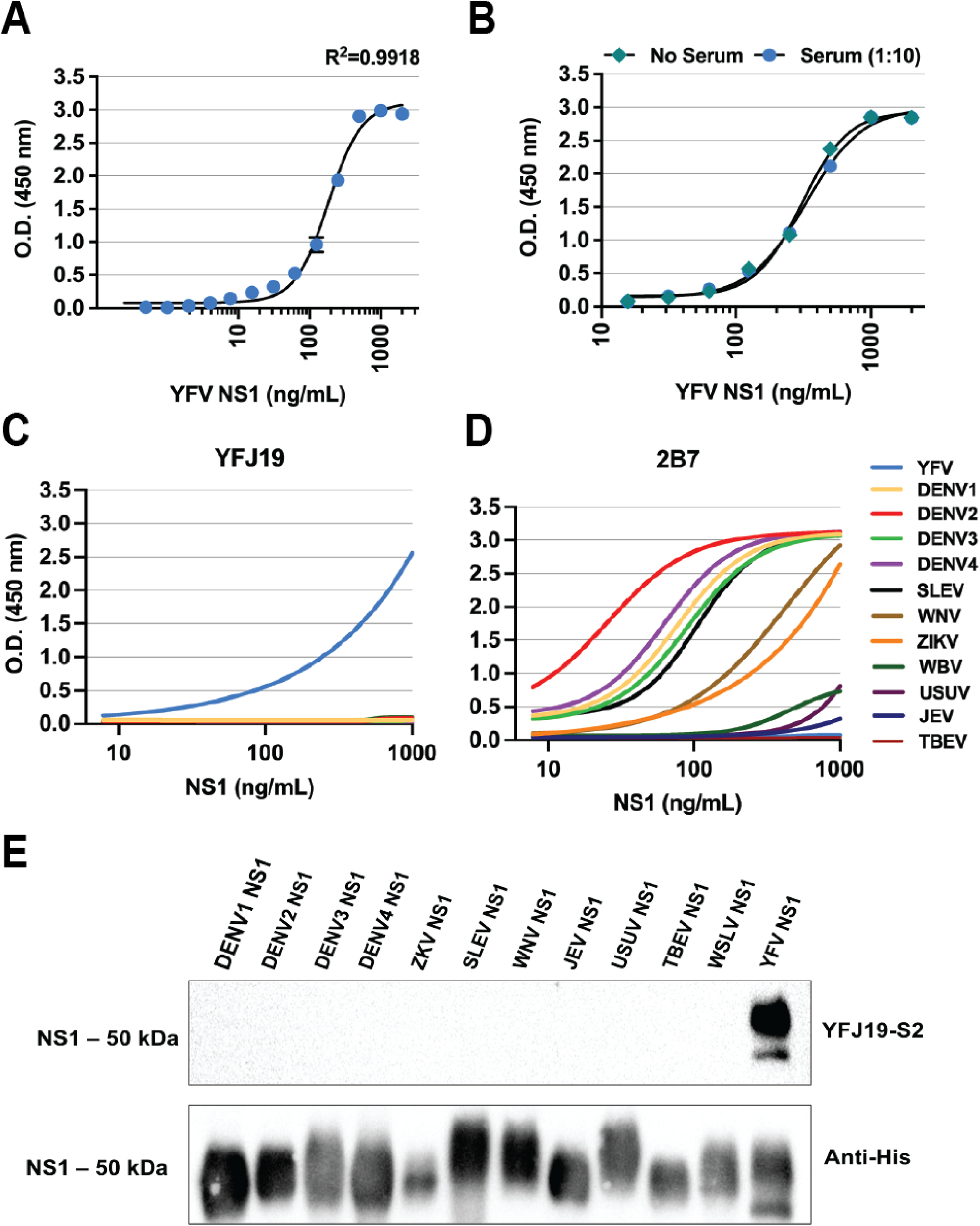
Development of quantitative YFV NS1 ELISA. (*A*) Sigmoidal standard curve of YFV NS1 capture ELISA performed with YFJ19 mAb and thirteen different concentrations of recombinant YFV NS1. (*B*) Comparison of standard curve of YFV NS1 diluted or not in normal human serum (1:10). (*C*) Specificity of mAb YFJ19 by direct ELISA performed with NS1 (5 ug/mL) from 12 different flaviviruses as follows: YFV; dengue virus serotypes 1 (DENV1), 2 (DENV2), 3 (DENV2), and 4 (DENV4); Saint Louis encephalitis virus (SLEV); West Nile virus (WNV); Zika virus (ZIKV); Wesselsbron virus (WBV); Usutu virus (USUV); Japanese encephalitis virus (JEV); Tick-borne encephalitis virus (TBEV). (*D*) Cross-reactivity an anti-flavivirus NS1 mAb (2B7) by direct ELISA performed with NS1 (5 ug/mL) from the 12 different flaviviruses as in C. (*E*) Western blot analysis showing specificity of mAb YFJ19 for YFV NS1.

### NS1 serum levels are significantly increased in severe YF

Using our in-house ELISA, we found that YFV NS1 levels were significantly higher in the severe YF group (mean=118.8 ng/mL) compared to the non-severe group (mean=29.5 ng/mL) and to the control group (no NS1 detected) (Fig. 2A). We also observed trending higher levels of YFV NS1 when comparing the deceased to the surviving group (Fig. 2B). YFV NS1 serum levels were observed to be highest between 5-13 days post-symptoms onset (Fig. 2C).

**Fig. 2.**
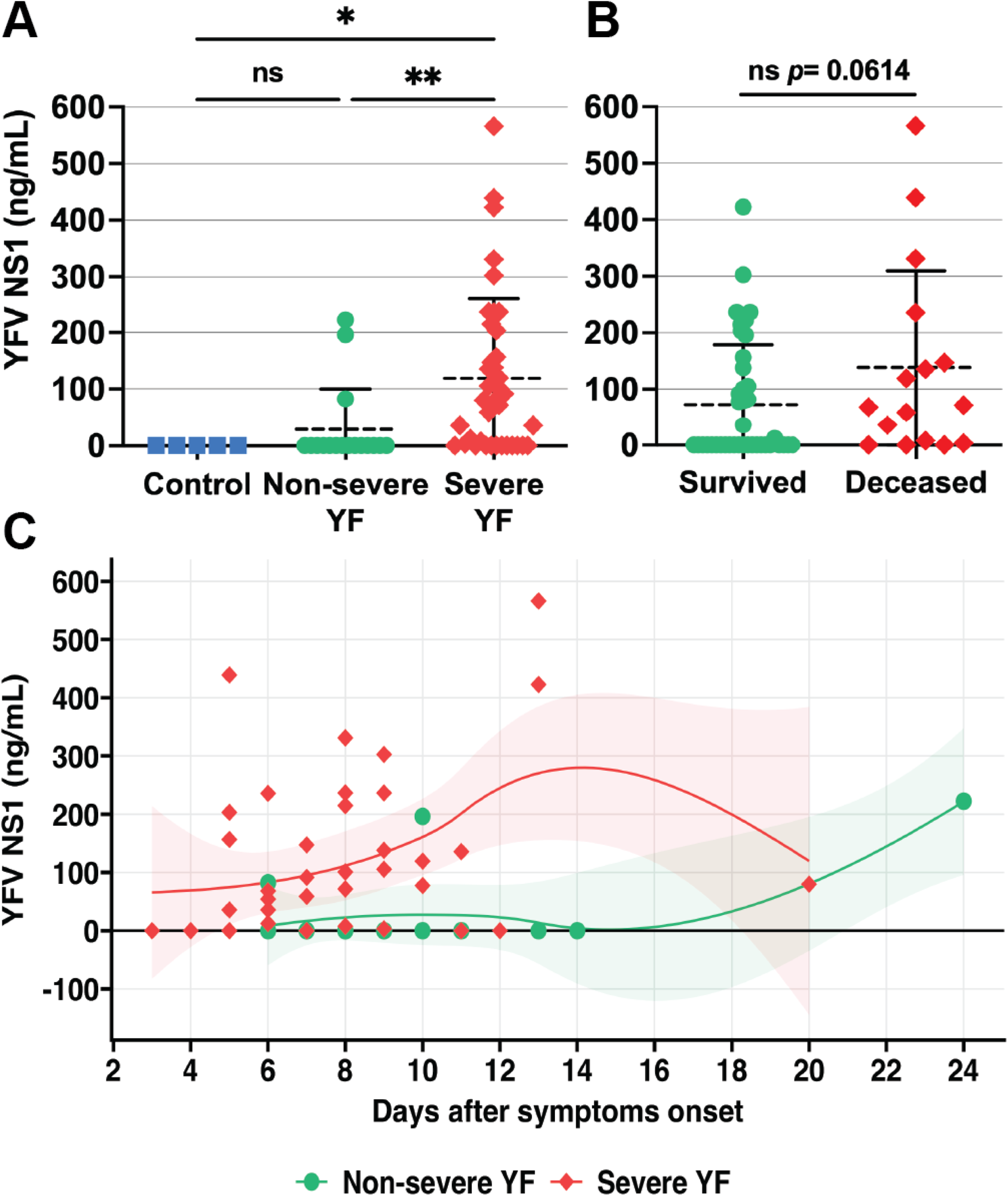
YFV NS1 serum levels in severe and non-severe YF patients. (*A*) YFV NS1 levels were determined using an in-house sandwich ELISA, as described in Materials and Methods. Studied individuals were classified in three different groups as described in Materials and Methods: 1) Severe YF (n=39); non-severe YF (n=16); controls (n=5): healthy individuals. (*B*) YFV NS1 levels in survivors and deceased YFV-infected groups. Mean ranks of groups were compared by Mann-Whitney test (significance level of 0.05). (*C*) YFV NS1 serum levels in individuals with acute YF according to days since symptom onset, visualized using a LOESS model.

### Serum levels of syndecan-1 correlate with disease severity in YF patients

Because sSDC-1 is a biomarker for endothelial injury in multiple diseases, we quantified sSDC-1 in the serum samples of our cohort. We found significantly increased sSDC-1 levels in severe vs. non-severe YF cases and significantly higher levels in both non-severe and severe YF groups when compared to healthy controls (Fig. 3A). Importantly, sSDC-1 levels were also significantly higher in individuals who succumbed compared to those who survived (Fig. 3B). These findings indicate that endothelial dysfunction is associated with disease severity in YFV-infected patients and that sSDC-1 can be used as a biomarker of YF disease severity. Similar to YFV NS1, sSDC-1 serum levels were highest 4-13 days after symptom onset (Fig. 3C). We also found that, as expected, sSDC-1 levels when compared to the same healthy controls as above were significantly increased in serum samples from dengue patients with diagnosed vascular leak obtained in our previous study (26) (Fig. 3D).

**Fig. 3.**
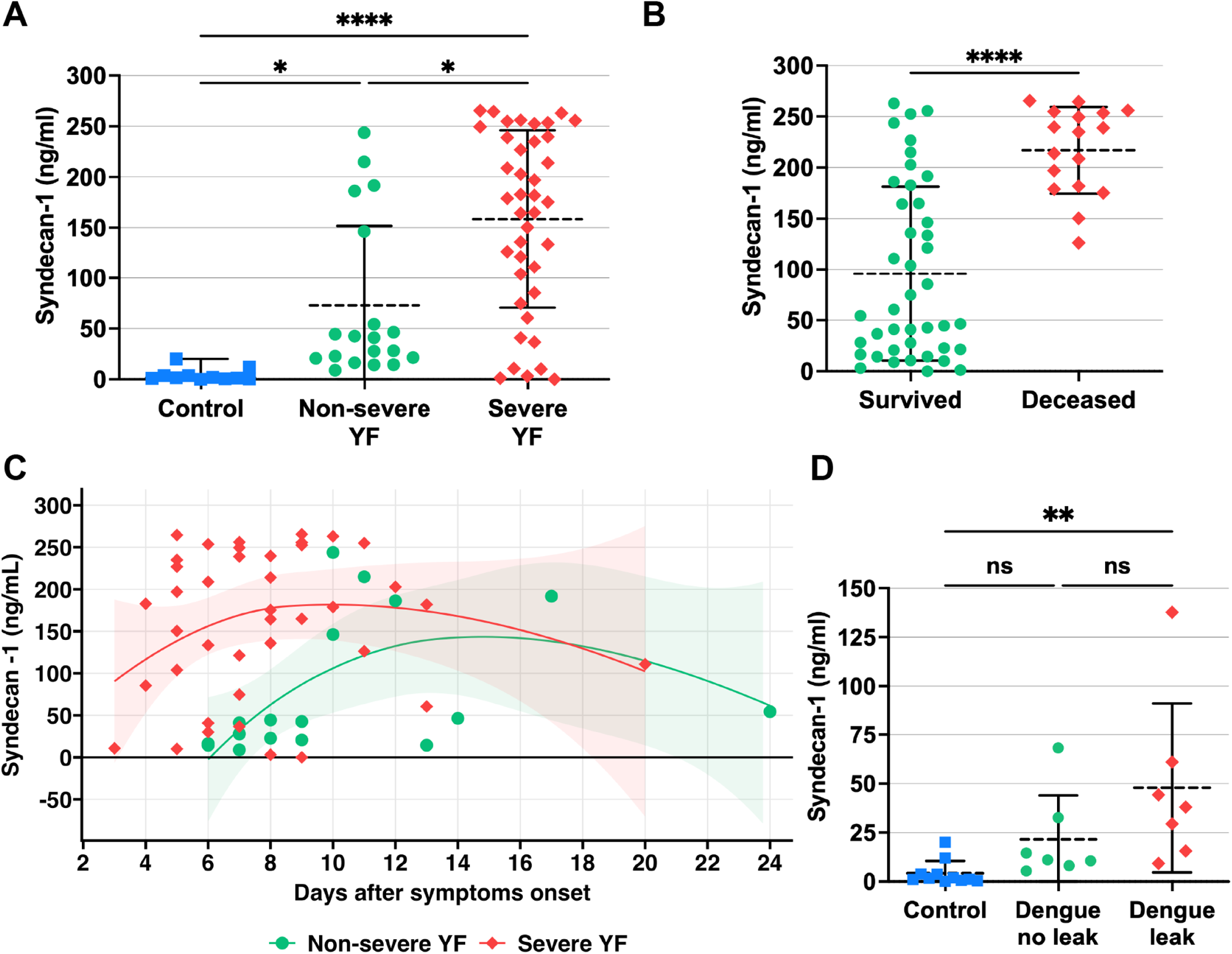
sSDC-1 levels in sera of individuals with acute YF and dengue. (*A*) The sSDC-1 levels in sera were determined by the Human sSDC-1 ELISA Kit. Studied individuals were classified in three different groups as described in Materials and Methods: 1) Severe YF (n=39); non-severe YF (n=18); controls (n=11): healthy individuals. (*B*) sSDC-1 serum levels in survivors and deceased YFV-infected groups. (*C*) Distribution and average (solid lines) of sSDC-1 serum levels in individuals with acute YF according to days since symptoms onset, visualized using a LOESS model. (*D*) sSDC-1 serum levels in individuals with acute DENV infection. Dengue no leak (n=7): individuals with acute dengue who did not display plasma leakage; Dengue leak (n=7): individuals with acute dengue who displayed plasma leakage; Controls (n=11): same healthy individuals shown in panel A. Mean ranks of sSDC-1 levels of YF or dengue groups were compared by Kruskal-Wallis + Dunn’s multiple comparisons test. Median of sSDC-1 levels in deceased and survived patients were compared Mann Whitney test (significance level of 0.05).

### Sera from patients acutely infected with YFV induce endothelial dysfunction *in vitro,* correlating with disease severity

To evaluate the capacity of serum from YF patients to mediate endothelial hyperpermeability, we evaluated the transendothelial electrical resistance (TEER) of human endothelial cells cultured in Transwell inserts and treated with sera from the different groups. We found that mean relative TEER values of severe and non-severe YF groups were reduced in comparison to medium-only or control groups, indicating that serum components were capable of inducing varying degrees of endothelial hyperpermeability according to disease severity (Fig. 4A). Comparing the area under the curve (AUC) of TEER nadirs, the severe YF group displayed significantly greater values when compared to both non-severe YF and control groups (Fig. 4B). Similarly, recombinant YFV NS1, used as positive control, induced endothelial dysfunction resulting in reduction of relative TEER values (Fig. 4A, B). The TEER curves for each sample are shown in Supplementary Fig. 2.

**Fig. 4.**
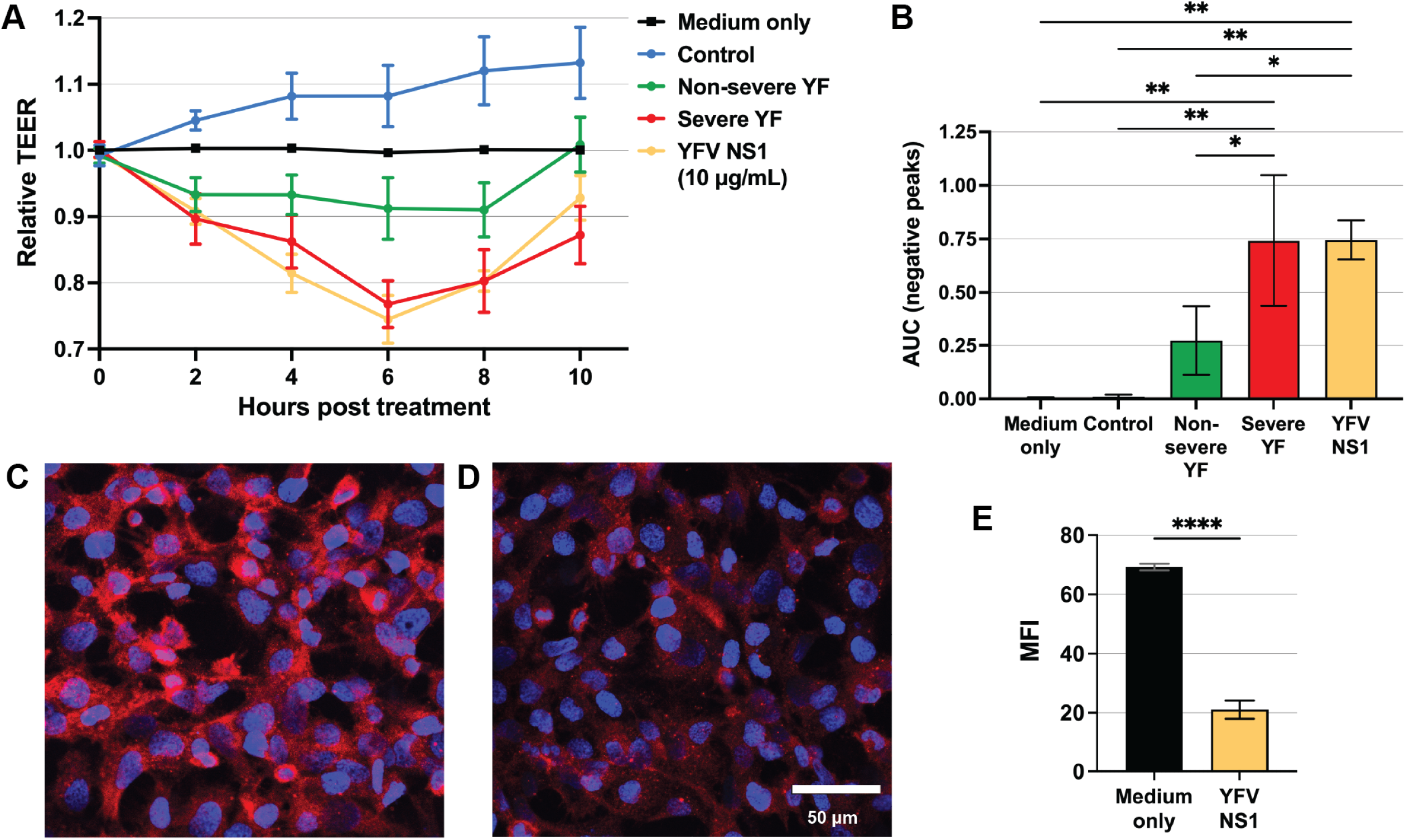
Relative TEER of human endothelial cells treated with acute YF human serum samples and IFA of SDC-1 after NS1 treatment. (*A*) Confluent monolayers of human endothelial cells cultured in Transwell inserts were treated or not with 10% serum from three different groups: 1) Severe YF (n=24); Non-severe YF (n=10); Controls (n=11): healthy individuals. YFV NS1 (10 ug/mL) was used as positive control. The transendothelial electrical resistance (TEER) was measured from 2 to 10h post-treatment. Graph shows mean ± SD of relative TEER for each treatment group. (*B*) Area under the curve (AUC) calculation for the different treatments. Mean values of AUC for each group were compared to untreated medium-only control group using one-way ANOVA + Tukey’s test. *(C, D)* Human endothelial cell monolayers grown in gelatin-coated coverslips were treated with *(C)* medium only or *(D)* 10 ug/mL of YFV NS1 for 3h. After fixation, cell surface SDC-1 was stained in red and nuclei in blue. *(E)* Quantification of SDC-1 protein on to the cells surface was expressed as mean fluorescence intensity (MFI) as shown in panels C and D. NS1 treatment was compared to medium-only treatment by t-test. Asterisks indicate significant difference with *p*<0.05.

### YFV NS1 induces sSDC-1 shedding in endothelial cells

To evaluate the capacity of YFV NS1 to serve as a direct trigger of endothelial dysfunction, we measured the effect of NS1 on SDC-1 levels on a monolayer of human endothelial cells. Consistent with our observed correlation between YFV NS1 and sSDC-1 in clinical samples, we found that YFV NS1 treatment of endothelial cells resulted in a significant reduction of cell surface-bound SDC-1 relative to medium only control (Fig. 4C, D, E). These data support a direct role for YFV NS1 as a contributing factor to endothelial dysfunction in YF patients.

### TEER, sSDC-1, and YFV NS1 serum levels correlate with parameters of severe YF

We next performed a correlation matrix analysis of patient data and results regarding TEER, sSDC-1 and YFV NS1 levels (Fig. 5A, with significant [p<0.05, r>0.35] correlations indicated with asterisks). Importantly, we found that YFV NS1 levels correlated with sSDC-1 levels, TEER values, neutrophil count, hematocrit (Ht), and IB (Fig. 5A, B, C). Further, sSDC-1 levels correlated significantly with YFV NS1, TEER values, gender, hospitalization, viral load, parameters of liver impairment (AST, ALT, creatinine, TB, DB, and IB), kidney dysfunction (creatinine), coagulopathy (fibrinogen, PT/INR, aPTT), and death (Fig. 5A, D). TEER values correlated with YFV NS1, sSDC-1, gender, creatinine, TB, DB, and IB (Fig. 5A, C, D). Thus, these findings show that TEER values and levels of YFV NS1 and sSDC-1 correlated with each other and with clinical laboratory parameters of disease severity, suggesting a potential link between YFV NS1 and endothelial dysfunction, resulting in increased shedding of sSDC1.

**Fig. 5.**
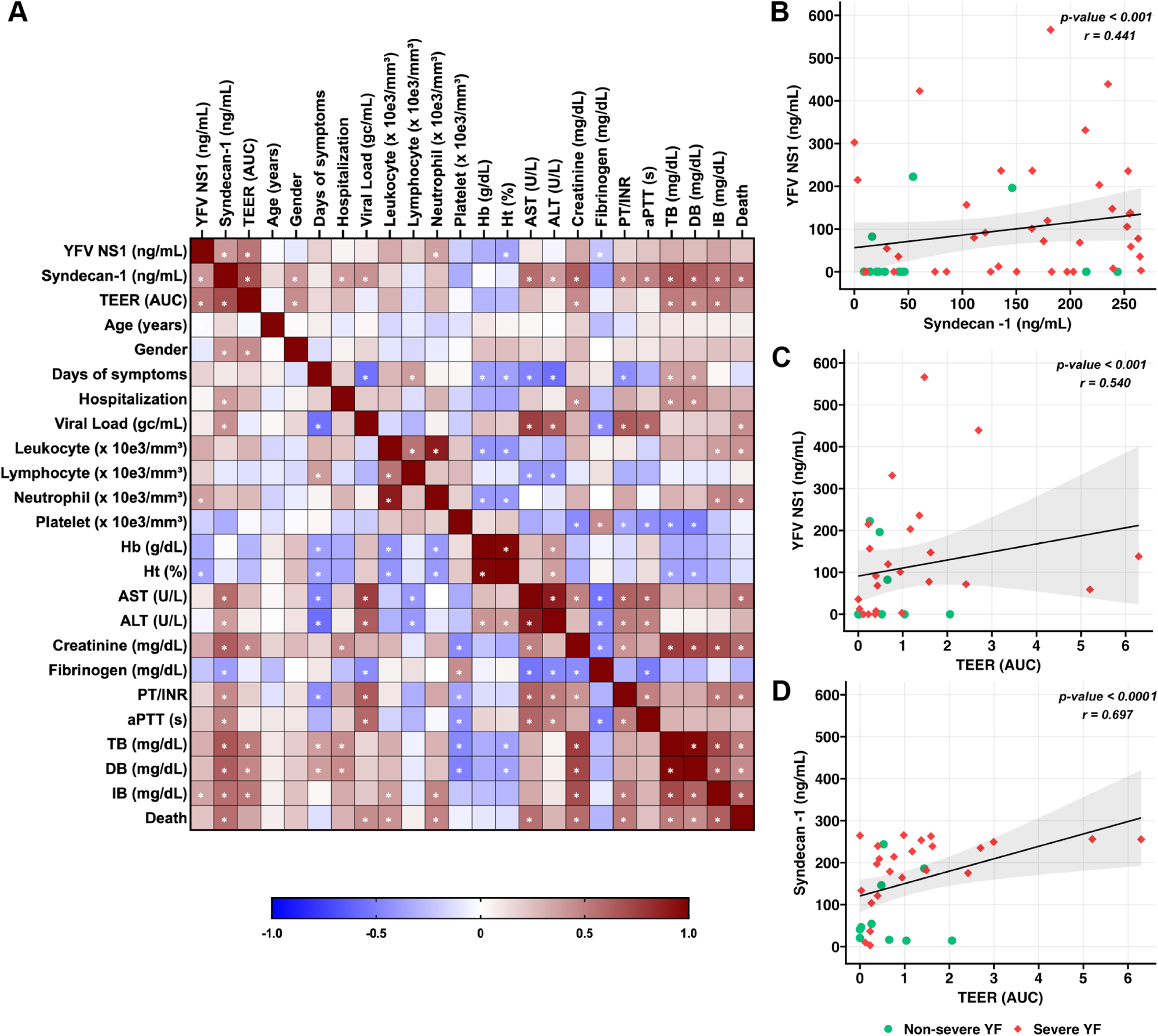
Correlation analysis of clinical and laboratory data of study participants during acute YF. (*A*) Correlation matrix analyzed by Spearman test using a linear model. Asterisks indicate significant correlations (p<0.05, Spearman r>0.35). (*B*) Correlation between serum levels of YFV NS1 and sSDC-1. (*C*) Correlation between serum levels of YFV NS1 and TEER AUC values. (D) Correlation between serum levels of sSDC-1 and TEER AUC values. Abbreviations: TEER, transendothelial electrical resistance; AUC, area under the curve; Hb, hemoglobin; Ht, hematocrit; AST, aspartate transaminase; ALT, alanine aminotransferase; TPI/INR thrombin potential index/international normalized ratio; aPTT, activated partial thromboplastin time; TB, total bilirubin; DB, direct bilirubin; IB, indirect bilirubin.

## Discussion

Our findings in this study indicate that in humans, YFV NS1 may directly target the endothelium, triggering breakdown of the glycocalyx via enzymatic activation, consistent with our previous report using *in vitro* and *in vivo* models (8). Serum levels of both NS1 and sSDC-1 were correlated with several clinical laboratory parameters associated with YF disease, providing evidence that YFV NS1 and sSDC-1 may be linked to severe YF disease manifestations. When placing human endothelial cells in contact with sera from the different groups, we observed a significant increase in permeability with the severe as compared to non-severe and healthy control groups. We also confirmed *in vitro* by IFA that YFV NS1 induces shedding of sSDC-1 from human endothelial cells. Our results highlight the secretion of YFV NS1 from infected cells and endothelial glycocalyx degradation, as measured by sSDC-1 shedding, as prominent characteristics of YF pathogenesis in humans and suggest that levels of NS1 and/or levels of sSDC-1 may help assess risk of developing severe illness.

Flavivirus NS1 is an important virulence factor in facilitating flavivirus pathogenesis by triggering enzymatic disruption of the glycocalyx as well as mediating the breakdown of intercellular junctional complexes of endothelial cells, both pathways contributing to tissue-specific vascular leak (8, 9, 27). We have previously shown that YFV NS1 treatment induces significant hyperpermeability in human endothelial cells and in mice (8). Our data here shows that severe YF cases displayed higher levels of YFV NS1 as early as 5 days post-disease onset, supporting the concept that production of NS1 by YFV-infected cells may contribute to YF disease. Thus, measuring the serum levels NS1 could potentially provide a clinical diagnostic/prognostic marker for YF.

SDC-1 is a heparan sulfate proteoglycan expressed on endothelial cells and an established marker of inflammatory disease and glycocalyx damage (28). Shedding of endothelial glycocalyx components, such as sSDC-1, is a known result of endothelial dysfunction, and increased serum levels of sSDC-1 have been confirmed in patients with diseases such as dengue (16, 25, 29, 30), COVID-19 (24), septic shock (31), and cancer (32, 33). Here, we show that higher levels of sSDC-1 correlate with severe YF; thus, sSDC-1 may also serve as a biomarker for severe YF disease.

YF is a systemic disease, but the pathology is primarily targeted to the liver. A recent study highlighted endothelial activation, showing immunohistochemical analysis of endothelial tissues in the hepatic parenchyma of YF-positive subjects, with increased expression of adhesion molecules (34). This study suggested that YFV induces endothelial activation, stimulating the rolling, recruitment, and migration of immune cells that contribute to inflammatory processes in the liver of fatal cases. Our current results highlight secretion of YFV NS1 into the bloodstream, where it presumably interacts with the endothelium, activating glycocalyx degradation and SDC-1 shedding that leads to endothelial hyperpermeability and possibly contributing to the hepatic and systemic inflammatory response.

In a previous study by Kallas et al. (2019), older age, male sex, higher leukocyte and neutrophil counts (>4000 cells/ mL), AST, bilirubin, creatinine, and viral load (>5.1 log10 copies/mL), as well as prolonged prothrombin time, were associated with increased mortality in patients with YF (2). Thus, we used these parameters to classify severe cases of YF in our current study, which utilized samples obtained from the same cohort. In a more recent study, hyperimmune activation and perturbation of the gut microbiome related to heightened levels of microbial translocation were also found to be associated with severe outcome in these patients (35). Moreover, coagulopathy in YF patients from the same cohort and in YFV-infected macaques was not only associated with defects in clotting factor synthesis due to hepatocyte infection, but also with coagulation factor consumption, shown by increased concentrations of plasma D-dimer (36).

Antigenic cross-reactivity during diagnosis of flaviviral diseases is a major challenge, particularly in endemic settings where multiple flaviviruses co-circulate (37). Therefore, assays measuring NS1 in sera must be highly specific to avoid incorrect diagnosis. The presence of NS1 is widely used as an early diagnostic marker for flavivirus infections, especially dengue (38). We produced and tested multiple novel anti-YFV NS1 mAbs when developing our YFV NS1 capture ELISA, aiming for minimal cross-reactivity to other flavivirus NS1 proteins while maintaining the highest sensitivity possible. Because our newly developed YFV NS1 ELISA is highly specific and can detect low concentrations of NS1 in supernatants or sera, it could be used in research and clinical settings, including differential diagnosis of flaviviruses at lower cost compared to molecular assays, with additional potential for prognosis.

Taken together, our study identifies YFV NS1 and sSDC-1 as correlates of YF disease severity and endothelial dysfunction. Our work suggests that endothelial dysfunction is a major clinical manifestation of human YF, as in dengue, and that circulating NS1 may contribute to this pathogenesis, as has been observed *in vitro* and in murine models. Further, our development of a highly specific YFV NS1 capture ELISA can serve as a proof-of-concept for the development of low-cost NS1-based diagnostics of YF and perhaps be used to predict YF disease progression in humans.

## Methods

### Study participants

This study utilized samples from patients with YF cases confirmed by detection of YFV genomic RNA in plasma and/or autopsy tissues by qRT-PCR, as previously described (2). Patients were enrolled at the Hospital das Clínicas, School of Medicine, University of São Paulo, Brazil, during an observational cohort study from January 2018 through February 2019 (2). Blood samples were collected at the time of admission after informed consent and before any treatment intervention or diagnostic procedure. Blood samples were used for clinical laboratory tests including determination of viral load; leukocyte and platelet counts; levels of hemoglobin, AST, ALT, creatinine, fibrinogen, and total, direct, and indirect bilirubin; and coagulation time. Cases were classified in three groups, as follows: i) non-severe (N=18): individuals who presented viral load <10^5^ genomic copies/mL, neutrophil count <4000/mL, AST <3500 U/L, creatinine <2.36 mg/mL, and IB <0.64 mg/dL and who recovered; ii) severe (N=39): individuals who presented one or more of the following criteria: viral load ≥10^5^ genomic copies/mL; neutrophil count ≥4000/mL, AST ≥3500 U/L, creatinine ≥2.36 mg/mL, IB ≥0.64 mg/dL, and/or death; iii) control (N=11): healthy individuals. The study was approved by the Institutional Review Board of the University of São Paulo (approval # CAAE: 59542216.3.1001.0068).

### Hybridoma production and ELISA and Western blot analysis

BALB/c mice were immunized intraperitoneally three times with 10 µg of recombinant YFV NS1 (Strain 17D, Native Antigen) diluted 1:1 in Sigma adjuvant system, and a fourth immunization with YFV NS1 alone. Splenocytes were fused with A1 myeloma cells, and hybridomas were selected on hypoxanthine-aminopterin-thymidine medium and screened with a YFV NS1 antigen-coat ELISA. In brief, Nunc MaxiSorp ELISA plates (Thermo Scientific) were coated overnight with 0.5 µg/mL of YFV NS1 in PBS and blocked the next day for 1 hour (h) using PBS containing 5% nonfat dry milk, washed twice with PBS, and incubated with hybridoma supernatant for 1h at room temperature. Plates were washed 3X with PBS-Tween20 0.01% (PBS-T) and 2X with PBS, and HRP-conjugated secondary antibody diluted in PBS-BSA 1% was added for 1h. After washing with PBS-T, 3,3′,5,5′-Tetramethylbenzidine (TMB) liquid substrate (Sigma) was added and left to develop for 10 minutes. Plates were read at OD_450_ nm using a BioTek/Agilent microplate reader. The mAbs with the highest OD value and shortest growth doubling time were selected for expansion and purification. Cells from ELISA-positive wells were sub-cloned and expanded before being tested by ELISA for YFV specificity using 12 recombinant flavivirus NS1 proteins purchased from Native Antigen Company: YFV; dengue virus serotypes 1 (DENV1), 2 (DENV2), 3 (DENV2), and 4 (DENV4); Saint Louis encephalitis virus (SLEV); West Nile virus (WNV); Zika virus (ZIKV); Wesselsbron virus (WBV); Usutu virus (USUV); Japanese encephalitis virus (JEV); and tick-borne encephalitis virus (TBEV) (31).

The YFJ19 mAb was tested by Western blot for specificity to YFV NS1. The same 12 recombinant flavivirus NS1 proteins used above were separated on a 10% polyacrylamide gel and transferred onto nitrocellulose membranes. Membranes were incubated overnight with 7 mL of hybridoma supernatant or 3.5 µL of mouse anti-His mAb (Abcam) as a control in PBS-T containing 5% nonfat dry milk. After antibody incubation, membranes were washed 4X with PBS-T and then probed with anti-mouse secondary antibodies conjugated to horseradish peroxidase (HRP; Biolegend) for 1h. Membranes were then washed 4X with PBS-T, developed using ECL reagents, imaged on a ChemiDoc system, and analyzed using Image Lab software (Bio-Rad).

### mAb purification

Hybridoma supernatant in batches of 400 mL were filtered using a 0.22 µm pore size bottle-top vacuum filter (Corning) to remove cell debris. Buffers were developed based on the manufacturer’s recommendations (Cytiva Protein G Sepharose 4 Affinity Chromatography Handbook). Supernatant was diluted 1:1 with binding buffer (20 mM H_2_NaO_4_P·H_2_O, pH 7.0). Protein G resin (2.5 mL, Cytiva Protein G SepharoseTM 4 Fast Flow) was added to a 1.0 x 10 cm Econo-Column (Bio-Rad) and washed twice with 10 mL of binding buffer. Supernatant/binding buffer solution was added to a Econo-Column Reservoir (Bio-Rad) attached to the Econo-Column and allowed to gravity flow through the resin. After flow-through was collected, the resin was washed twice with 10 mL of binding buffer to remove supernatant. To elute the bound mAbs, 6 mL of elution buffer (0.1 M glycine buffer, pH 2.5-3.0) was added to the column. Elution fractions were collected in 1-mL fractions and diluted 1:10 with neutralizing buffer (1 M TrisHCl, pH 9.0). mAb concentration was calculated using a Nanodrop spectrophotometer, and the top yields were selected and pooled. Purified mAbs were dialyzed using a Slide-A-Lyzer 10K (Fisher) over 48h at 4°C with two separate exchanges of PBS buffer.

### YFV NS1 capture ELISA

ELISA plates were coated with 5 µg/mL of capture mAb in 50 µL PBS/well and incubated at 4°C overnight. The next day, plates were washed once with PBS and blocked with PBS-BSA 3% (100 µL) and incubated for 1h at room temperature. The plate was then washed twice with PBS-T, and the serum or recombinant NS1 was diluted in PBS-BSA 1% (50 µL) before being added to the plate and incubated for 1h at room temperature. Plates were washed 4X with PBS-T, and the biotinylated detecting mAb (Pierce Antibody Biotinylation Kit for IP, ThermoFisher) was diluted in PBS-BSA 1% (50 µL) and added to the plate to incubate for 1h at room temperature. The plate was then washed 4X with PBS-T. HRP-streptavidin (Jackson Immuno) diluted in PBS-BSA (50 µL) was then added to the plate and incubated at room temperature for 1h. After incubation, the plate was washed 4X with PBS-T and 1X with PBS. The plate was then developed with TMB substrate (100 µL; Sigma) for ∼15 minutes. The enzymatic reaction was interrupted using 2N H_2_SO_4_, and the plate was read at OD_450_ nm. The concentrations of NS1 in sera were interpolated using a standard curve of recombinant NS1 ranging from 1 to 1000 ng/mL; the limit of detection for this assay was 2 ng/mL.

### Determination of sSDC-1 levels in human serum

The amount of sSDC-1 in serum was determined using the human SDC-1 DuoSet ELISA Kit (DY2780, R&D Systems). Briefly, 96-well ELISA microplates were coated with 80 ng/well of goat anti-human SDC-1 capture antibody overnight at room temperature. After washing and blocking with 1% bovine serum albumin (BSA), plates were incubated with samples diluted 1:25 or recombinant human SDC-1 standards for 2h. After washing, plates were incubated with 5 ng/well of biotinylated goat anti-human SDC-1 detection antibody. After 2h, HRP-streptavidin was added for signal detection with TMB substrate. The OD_450_ with a correction at OD_550_ was determined using a microplate reader. sSDC-1 levels were determined by interpolation analysis of standard curves with four-parameter logistic regression.

### Evaluation of endothelial barrier function *in vitro*

To evaluate the putative effects of serum from acutely YFV-infected patients on endothelial barrier function, we used the TEER assay as described previously (26, 27). In brief, human umbilical vein endothelial cells, kindly donated by Dr. Miriam Fonseca-Alaniz (Instituto do Coração, InCor, University of São Paulo, Brazil) were seeded (6×10^4^ cells/well) in Transwell polycarbonate membrane inserts (0.4 μm pore, 6.5 mm diameter; Corning Inc.) in endothelial cell growth basal medium 2 supplemented with an Endothelial Cell Growth Medium-2 (EGM-2TM) supplemental bullet kit (Lonza). After 72h of incubation at 37°C and 5% CO_2_, cells were treated with human sera (10% final vol/vol concentration) obtained from YFV-positive severe and non-severe patients or YFV-negative blood donors (healthy controls). TEER values, expressed in Ohms (Ω), were collected at sequential 2-h time-points 2-10h following treatments using an Epithelial Volt Ohm Meter (EVOM) with a “chopstick” electrode (World Precision Instruments). Resistance of inserts with no cells (blank) and inserts with cells (untreated) containing medium alone, were used to calculate relative TEER as a ratio of the corrected resistance values as (Ω experimental condition - Ω blank)/(Ω untreated - Ω blank). Recombinant YFV NS1 (Native Antigen Co.) at 10 µg/mL was used as positive control.

### Measurement of SDC-1 levels on the surface of human endothelial cells by immunofluorescence assay (IFA)

HPMEC (1×10^5^ cells) were seeded on gelatin-coated coverslips and allowed to grow until full confluency was attained (approximately 3 days). On the day of the experiment, cells were treated or not with 10 μg/mL of YFV NS1 protein and incubated for 3h at 37°C. After incubation, cells were washed, fixed with 4% paraformaldehyde (PFA), and stained overnight with 2 ug/mL of the rabbit anti-SDC-1 IgG antibody (Abcam, ab188861). After washing, secondary staining was performed by adding 2 ug/mL of donkey anti-rabbit IgG conjugated to Alexa Fluor 647 (Abcam, ab150075) for 4h. Nuclei were stained using Hoechst (ImmunoChemistry Technologies). Mounted slides were imaged on a Zeiss LSM 710 Axio Observer fluorescence microscope (CRL Molecular Imaging Center, UC Berkeley). Images acquired using the Zen 2010 software (Zeiss, Jena, Germany) were processed and analyzed with ImageJ software (39). Mean fluorescence intensity (MFI) values for SDC-1 staining were obtained from individual RGB-grayscale-transformed images (n = 3).

### Statistics

ELISA values were modeled using 4-parametric logistic regression, which follows a sigmoidal distribution to represent the optical density range (40). Spearman’s rank correlation was used to determine the relationship between two non-parametric continuous variables (41). Linear models were used to visualize the relationship between continuous values of relevant biomarkers (42). Locally estimated scatterplot smoothing (LOESS) models were used to visualize the trend of relevant biomarkers over days since symptom onset (43). LOESS models were implemented using a span = 1. The span was determined based on the best visualization that accounts for the low sample size and large gaps between the sample days. Data were analyzed using R language version 4.1.1 within the RStudio (2021.09.0, Build 351) integrated development environment. Plots were visualized using the base R graphics and ggplot2 (v3.3.5) package. Statistical tests used in this study include ANOVA analysis with multiple comparisons test as well as t-tests, as indicated in the figure legends. Resultant *p*-values from the above statistical tests are displayed as ns, not significant (*p*≥0.05) or with asterisks as follows: \**p*<0.05, \*\**p*<0.01, \*\*\**p*<0.001, or \*\*\*\**p*<0.0001. All statistics not indicated are not significant.

## Supporting information

Supplemental Tramontini

## Data Availability

All data produced in the present study are available upon reasonable request to the authors

## Acknowledgments

This study was supported by NIH grants, R01 AI24493 (E.H.) and R01 AI168003 (E.H.), and by São Paulo Research Foundation-FAPESP (Project #2013/01690-0 to ECS and Scholarships #2013/01702-9 and 2017/16627-3 to FTGS). SBB was supported in part as an Open Philanthropy Awardee of the Life Sciences Research Foundation.

## Notes

### Competing Interest Statement

The authors have declared no competing interest.

### Funding Statement

This study was supported by NIH grants, R01 AI24493 (E.H.) and R01 AI168003 (E.H.), and by Sao Paulo Research Foundation-FAPESP (Project #2013/01690-0 to ECS and Scholarships #2013/01702-9 and 2017/16627-3 to FTGS). SBB was supported in part as an Open Philanthropy Awardee of the Life Sciences Research Foundation.

### Author Declarations

The Institutional Review Board of the University of Sao Paulo, gave ethical approval for this work. Approval # CAAE: 59542216.3.1001.0068.

## References

1. M. U. Kraemer, et al., Spread of yellow fever virus outbreak in Angola and the Democratic Republic of the Congo 2015–16: a modelling study. Lancet Infect. Dis. 17, 330–338 (2017).

2. E. G. Kallas, et al., Predictors of mortality in patients with yellow fever: an observational cohort study. Lancet Infect. Dis. 19, 750–758 (2019).

3. C. L. Gardner, K. D. Ryman, Yellow fever: a reemerging threat. Clin. Lab. Med. 30, 237–260 (2010).

4. D. R. Glasner, H. Puerta-Guardo, P. R. Beatty, E. Harris, The good, the bad, and the shocking: the multiple roles of dengue virus nonstructural protein 1 in protection and pathogenesis. Annu. Rev. Virol. 5, 227–253 (2018).

5. D. H. Libraty, et al., High circulating levels of the dengue virus nonstructural protein NS1 early in dengue illness correlate with the development of dengue hemorrhagic fever. J. Infect. Dis. 186, 1165–1168 (2002).

6. S. A. Paranavitane, et al., Dengue NS1 antigen as a marker of severe clinical disease. BMC Infect. Dis. 14, 570 (2014).

7. H. T. L. Duyen, et al., Kinetics of plasma viremia and soluble nonstructural protein 1 concentrations in dengue: differential effects according to serotype and immune status. J. Infect. Dis. 203, 1292–1300 (2011).

8. H. Puerta-Guardo, et al., Flavivirus NS1 Triggers Tissue-Specific Vascular Endothelial Dysfunction Reflecting Disease Tropism. Cell Rep. 26, 1598–1613.e8 (2019).

9. H. Puerta-Guardo, et al., Flavivirus NS1 Triggers Tissue-Specific Disassembly of Intercellular Junctions Leading to Barrier Dysfunction and Vascular Leak in a GSK-3β-Dependent Manner. Pathogens 11, 615 (2022).

10. P. Pan, et al., DENV NS1 and MMP-9 cooperate to induce vascular leakage by altering endothelial cell adhesion and tight junction. PLoS Pathog. 17, e1008603 (2021).

11. A. W. Wessel, et al., Levels of circulating NS1 impact West Nile virus spread to the brain. J. Virol. 95, e00844–21 (2021).

12. L. Hui, et al., Matrix metalloproteinase 9 facilitates Zika virus invasion of the testis by modulating the integrity of the blood-testis barrier. PLoS Pathog. 16, e1008509 (2020).

13. C.-Y. Lin, et al., High levels of serum hyaluronan is an early predictor of dengue warning signs and perturbs vascular integrity. EBioMedicine 48, 425–441 (2019).

14. D. A. Espinosa, et al., Increased serum sialic acid is associated with morbidity and mortality in a murine model of dengue disease. J. Gen. Virol. 100, 1515–1522 (2019).

15. H. Puerta-Guardo, et al., Zika Virus Nonstructural Protein 1 Disrupts Glycosaminoglycans and Causes Permeability in Developing Human Placentas. J. Infect. Dis. 221, 313–324 (2020).

16. V. Mariappan, S. Adikari, L. Shanmugam, J. M. Easow, A. B. Pillai, Expression dynamics of vascular endothelial markers: Endoglin and syndecan-1 in predicting dengue disease outcome. Transl. Res. 232, 121–141 (2021).

17. H.-R. Chen, et al., Macrophage migration inhibitory factor is critical for dengue NS1-induced endothelial glycocalyx degradation and hyperpermeability. PLoS Pathog. 14, e1007033 (2018).

18. T. H.-C. Tang, et al., Increased serum hyaluronic acid and heparan sulfate in dengue fever: association with plasma leakage and disease severity. Sci. Rep. 7, 1–9 (2017).

19. A. D. Theocharis, S. S. Skandalis, G. N. Tzanakakis, N. K. Karamanos, Proteoglycans in health and disease: novel roles for proteoglycans in malignancy and their pharmacological targeting. FEBS J. 277, 3904–3923 (2010).

20. L. Liu, M. Akkoyunlu, Circulating CD138 enhances disease progression by augmenting autoreactive antibody production in a mouse model of systemic lupus erythematosus. J. Biol. Chem. 297, 101053 (2021).

21. T. Manon-Jensen, Y. Itoh, J. R. Couchman, Proteoglycans in health and disease: the multiple roles of syndecan shedding. FEBS J. 277, 3876–3889 (2010).

22. K. Suzuki, et al., Serum syndecan-1 reflects organ dysfunction in critically ill patients. Sci. Rep. 11, 1–9 (2021).

23. R. Vollenberg, et al., Indications of persistent glycocalyx damage in convalescent COVID-19 patients: a prospective multicenter study and hypothesis. Viruses 13, 2324 (2021).

24. D. Zhang, et al., Syndecan-1, an indicator of endothelial glycocalyx degradation, predicts outcome of patients admitted to an ICU with COVID-19. Mol. Med. 27, 1–12 (2021).

25. S. Suwarto, R. T. Sasmono, R. Sinto, E. Ibrahim, M. Suryamin, Association of Endothelial Glycocalyx and Tight and Adherens Junctions With Severity of Plasma Leakage in Dengue Infection. J. Infect. Dis. 215, 992–999 (2017).

26. F. Tramontini Gomes de Sousa Cardozo, et al., Serum from dengue virus-infected patients with and without plasma leakage differentially affects endothelial cells barrier function in vitro. PloS One 12, e0178820 (2017).

27. S. B. Biering, et al., Structural basis for antibody inhibition of flavivirus NS1–triggered endothelial dysfunction. Science 371, 194–200 (2021).

28. S. Gaudette, D. Hughes, M. Boller, The endothelial glycocalyx: structure and function in health and critical illness. J. Vet. Emerg. Crit. Care 30, 117–134 (2020).

29. P. K. Lam, et al., Visual and biochemical evidence of glycocalyx disruption in human dengue infection, and association with plasma leakage severity. Front. Med. 7, 545813 (2020).

30. B. Buijsers, et al., Increased plasma heparanase activity and endothelial glycocalyx degradation in dengue patients is associated with plasma leakage. Front. Immunol. 12, Article number 759570 (2021).

31. A. Haynes III, et al., Syndecan 1 shedding contributes to Pseudomonas aeruginosa sepsis. Infect. Immun. 73, 7914–7921 (2005).

32. H. Joensuu, et al., Soluble syndecan-1 and serum basic fibroblast growth factor are new prognostic factors in lung cancer. Cancer Res. 62, 5210–5217 (2002).

33. Z. Malek-Hosseini, S. Jelodar, A. Talei, A. Ghaderi, M. Doroudchi, Elevated Syndecan-1 levels in the sera of patients with breast cancer correlate with tumor size. Breast Cancer 24, 742–747 (2017).

34. F. A. Olímpio, et al., Endothelium Activation during Severe Yellow Fever Triggers an Intense Cytokine-Mediated Inflammatory Response in the Liver Parenchyma. Pathog. Basel Switz. 11, 1–7 (2022).

35. A.-N. Pelletier, et al., Yellow fever disease severity is driven by an acute cytokine storm modulated by an interplay between the human gut microbiome and the metabolome. medRxiv, 2021.09.25.21264125 (2021).

36. A. L. Bailey, et al., Consumptive coagulopathy of severe yellow fever occurs independently of hepatocellular tropism and massive hepatic injury. Proc. Natl. Acad. Sci. 117, 32648–32656 (2020).

37. A. P. Rathore, A. L. St. John, Cross-reactive immunity among flaviviruses. Front. Immunol. 11, 334 (2020).

38. M. G. Guzman, et al., Multi-country evaluation of the sensitivity and specificity of two commercially-available NS1 ELISA assays for dengue diagnosis. PLoS Negl. Trop. Dis. 4, e811 (2010).

39. C. A. Schneider, W. S. Rasband, K. W. Eliceiri, NIH Image to ImageJ: 25 years of image analysis. Nat. Methods 9, 671–675 (2012).

40. C. Ritz, F. Baty, J. C. Streibig, D. Gerhard, Dose-response analysis using R. PloS One 10, e0146021 (2015).

41. W. J. Conover, Practical nonparametric statistics (John Wiley & Sons, 1999).

42. J. M. Chambers, A. Freeny, R. M. Heiberger, “Linear models” in Statistical Models, (J. M. Chambers and T. J. Hastie,Wadsworth & Brooks/Cole., 1992).

43. W. S. Cleveland, E. Grosse, W. M. Shyu, “Local regression models” in Statistical Models, (J. M. Chambers and T. J. Hastie,Wadsworth & Brooks/Cole., 1992).

