## Supplemental Tramontini for "Yellow fever disease severity and endothelial dysfunction are associated with elevated serum levels of viral NS1 protein and syndecan-1"

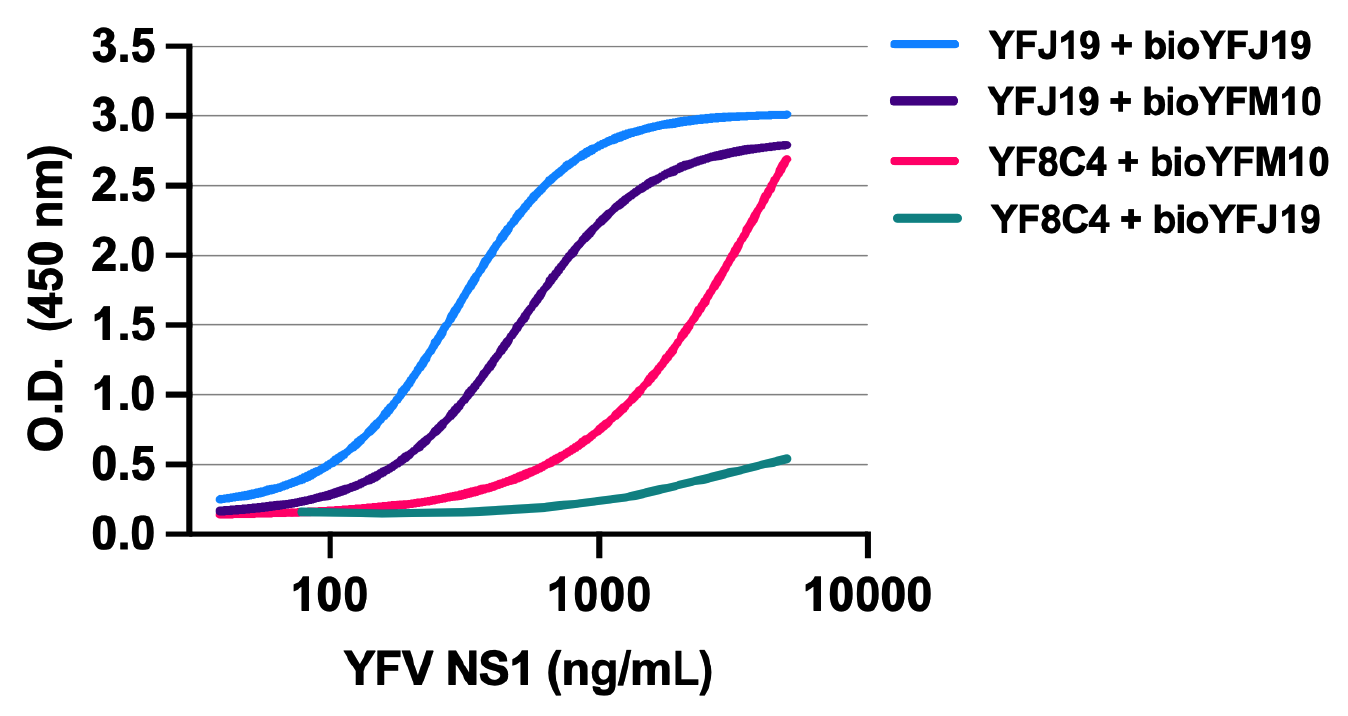

**Fig. S1. Comparison of standard curve of a capture YFV NS1 ELISA performed with different monoclonal antibodies combinations**. Standard curves of capture ELISA performed with different combinations of anti-YFV NS1 monoclonal antibodies for capture and biotinylated (Bio) antibodies for detection. Eight different concentrations (5000 to 39.06 ng/mL) of recombinant YFV NS1 were quantified showing that YFJ19 + bioYFJ19 was the best combination with higher signal.

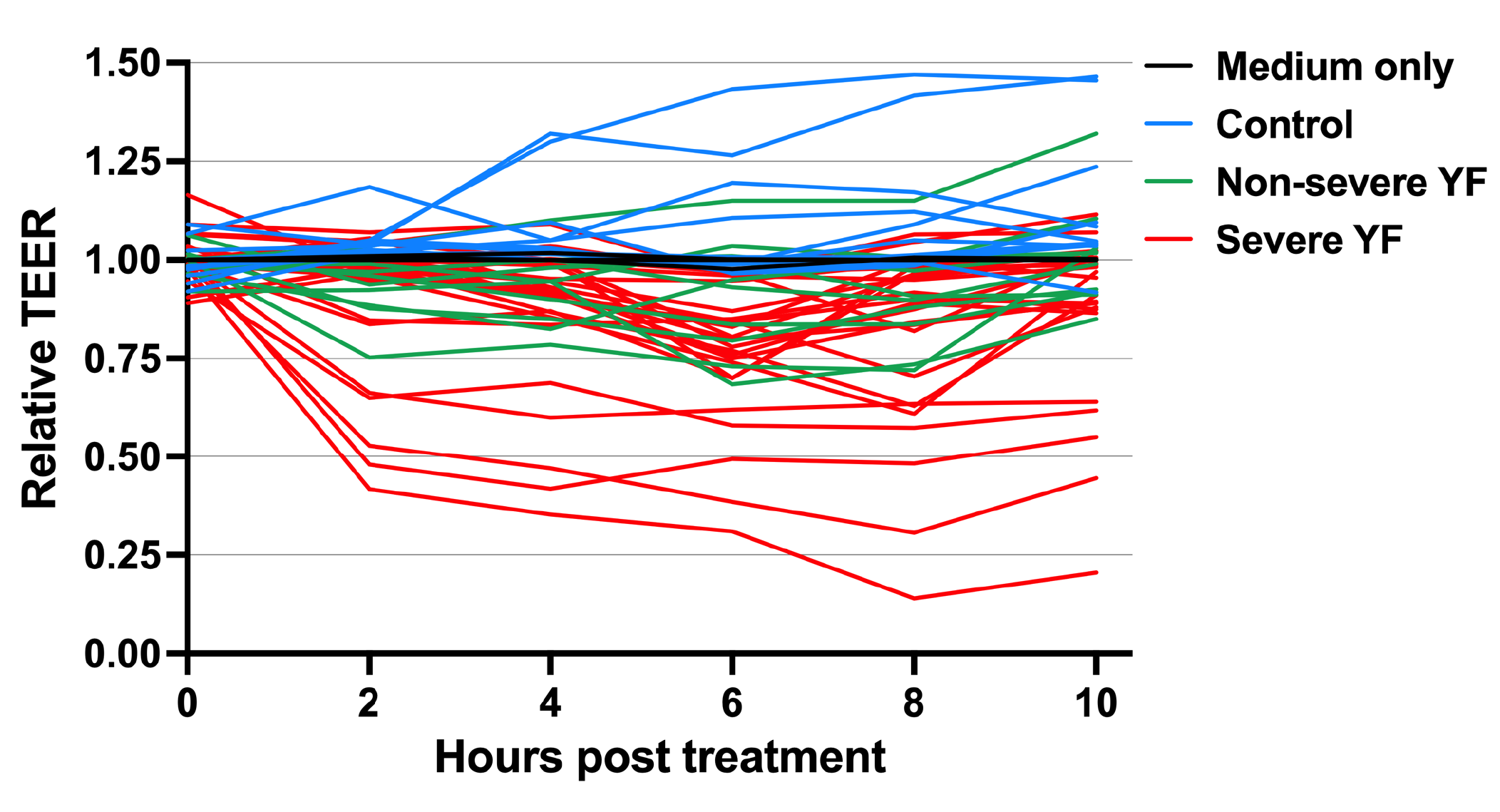

**Fig. S2. Relative TEER of human endothelial cells treated with acute YF human samples**. Confluent monolayers of human endothelial cells cultured in Transwell inserts were treated or not with 10% serum from three different groups: 1) Severe YF (n=24); Non severe YF (n=10); Control (n=11): healthy individuals. The transendothelial electrical resistance (TEER) was measured from 2 to 10h post-treatment. Graph shows data for each tested sample separately.
